# Prevalence and biochemical patterns of thyroid dysfunction among children and young adults with Type 1 Diabetes in Kenya: A multicentre cross-sectional study

**DOI:** 10.64898/2026.11.12.26367896

**Authors:** Wamalwa Phoebe, Kahssay Menbere, Martha Brenda Oeba, Winnie Mueni Saumu, Paul Laigong, Anjumanara Omar

## Abstract

**Background:** Thyroid dysfunction is a common endocrine comorbidity in type 1 diabetes (T1D), yet most evidence on its burden and biochemical patterns comes from higher-income settings. Data among African children and young adults with T1D remain limited.

**Objective:** To determine the prevalence and pattern of thyroid dysfunction and examine demographic and diabetes-related factors associated with thyroid dysfunction among children and young adults with T1D in Kenya.

**Methods:** We conducted a multicentre cross-sectional study among children and young adults with T1D aged ≤34 years attending four diabetes clinics in Kenya. Serum thyroid-stimulating hormone (TSH) and free thyroxine (FT4) were measured. Thyroid status was classified as euthyroidism (normal TSH and FT4), subclinical hypothyroidism (elevated TSH with normal FT4), overt hypothyroidism (elevated TSH with low FT4), subclinical hyperthyroidism (low TSH with normal FT4), overt hyperthyroidism (low TSH with elevated FT4), or discordant TSH–FT4 profiles that did not meet these predefined biochemical categories. Prevalence estimates with 95% confidence intervals (CI) were calculated, and multivariable logistic regression examined associations with age, sex and duration of diabetes.

**Results:** Among 244 participants with paired TSH and FT4 measurements, 77.5% (n=189) were euthyroid. Twenty-seven participants (11.1%; 95% CI 7.7–15.6%) had biochemically defined thyroid dysfunction: 18 (7.4%) had subclinical hypothyroidism, 5 (2.0%) overt hypothyroidism, and 4 (1.6%) subclinical hyperthyroidism. No overt hyperthyroidism was identified. An additional 28 participants (11.5%) had discordant TSH–FT4 profiles. Hypothyroidism accounted for 85.2% of biochemically defined thyroid dysfunction and was predominantly subclinical (78.3%). Age, sex and duration of diabetes were not significantly associated with thyroid dysfunction in adjusted analyses.

**Conclusion:** Thyroid dysfunction affected approximately one in nine Kenyan children and young adults with T1D and was predominantly hypothyroid, particularly subclinical hypothyroidism. Discordant TSH–FT4 profiles were also frequent, underscoring the importance of careful biochemical classification and confirmation of abnormal thyroid-function tests.

## 1. Introduction

Thyroid dysfunction is an important endocrine comorbidity in people living with type 1 diabetes (T1D), occurring more frequently than in the general population. ^1,4^ Hypothyroidism is the predominant thyroid disorder reported in T1D, with subclinical hypothyroidism occurring more commonly than overt disease, while hyperthyroidism is comparatively uncommon.^2,3^ A systematic review and meta-analysis involving 293,889 individuals with T1D estimated the pooled prevalence of hypothyroidism at 9.8% (95% CI 7.5–12.3%) and hyperthyroidism at 1.3% (95% CI 0.9–1.8%).^4^ Similarly, a recent analysis of 19,225 children with T1D enrolled in the international SWEET registry reported hypothyroidism in 9.9%, with considerable geographical variation^5^. This burden is substantially greater than that reported in children without T1D: population-based studies estimate subclinical hypothyroidism in approximately 1.7–2.9% of otherwise healthy children and adolescents^6,7^. Thyroid dysfunction may remain clinically silent, particularly in its subclinical form, but may adversely affect linear growth, lipid metabolism, body weight and glycaemic management^7,8^. These clinical consequences underpin recommendations for periodic thyroid-function assessment in children with T1D. The 2026 American Diabetes Association Standards of Care recommend measurement of thyroid-stimulating hormone (TSH) at diagnosis when metabolically stable, followed by repeat testing every 1–2 years when initial results are normal^1^.

The prevalence and biochemical pattern of thyroid dysfunction in T1D vary considerably between populations and may be influenced by age, sex, duration of diabetes, assay methodology, diagnostic thresholds, iodine exposure, geography and study design. ^5,8,9^ Iodine deficiency is unlikely to be a major explanation for thyroid-function differences in contemporary East African populations. In a multicentre study conducted in Kenya and Tanzania, urinary iodine concentrations were above thresholds for adequate iodine nutrition across population groups and were in the excessive range among school-age children, largely reflecting iodised salt exposure. Despite this, clinically significant thyroid dysfunction was uncommon in children and infants^8^. These findings suggest that thyroid-function patterns observed in East African populations should not automatically be attributed to iodine deficiency and support comparison with other iodine-sufficient populations, while recognising that genetic, environmental and methodological differences may still influence reported prevalence.

Evidence from sub-Saharan Africa remains limited and heterogeneous. In Uganda, Muhame and colleagues evaluated 69 children and adolescents aged 1–19 years with established T1D and found subclinical hypothyroidism in 7.3%, despite all participants being clinically euthyroid.^10^ In Upper Egypt, a controlled study of 94 children and adolescents with T1D reported subclinical hypothyroidism in 6.3% and overt hypothyroidism in 2.1%, compared with 2.1% and 0%, respectively, among age- and sex-matched controls.^11^ More recent Ethiopian data among diabetic populations have also demonstrated substantial thyroid dysfunction, although available studies have included mixed T1D and T2D populations and are therefore not directly comparable with paediatric T1D cohorts.^12^Taken together, these studies suggest that thyroid dysfunction is clinically relevant in African populations with diabetes, but estimates vary considerably according to population characteristics and study methodology.

Kenyan data are particularly sparse. To our knowledge, there are no published multicentre Kenyan studies characterising biochemical thyroid dysfunction among children and young people with T1D. Existing Kenyan thyroid literature has largely addressed iodine exposure, neonatal thyroid-function reference intervals and congenital hypothyroidism.^8,13^ We therefore sought to determine the prevalence and pattern of thyroid dysfunction and to examine its relationship with age, sex and duration of diabetes in a multicentre Kenyan cohort of children, adolescents and young adults with T1D.

## 2. Materials and Methods

### Study design and setting

We conducted a multicentre hospital-based cross-sectional study among children, adolescents and young adults living with T1D and receiving care at four diabetes clinics in Kenya. The clinics were selected to reflect variation in geography and level of healthcare delivery in Kenya. Kenyatta National Hospital (KNH) is a Level 6 national referral and teaching hospital in Nairobi with established paediatric endocrinology and adult diabetes services. Westlands Level 4 Hospital represents a lower-level urban facility within Nairobi County. Machakos Level 5 Hospital and Kajiado County Referral

Hospitals are county referral facilities located outside Nairobi and serve populations drawn largely from peri-urban, rural and semi-arid settings. Together, the four sites provided a diverse clinical setting for evaluating thyroid dysfunction among children and young adults with T1D (Figure 1).

**Figure 1.**
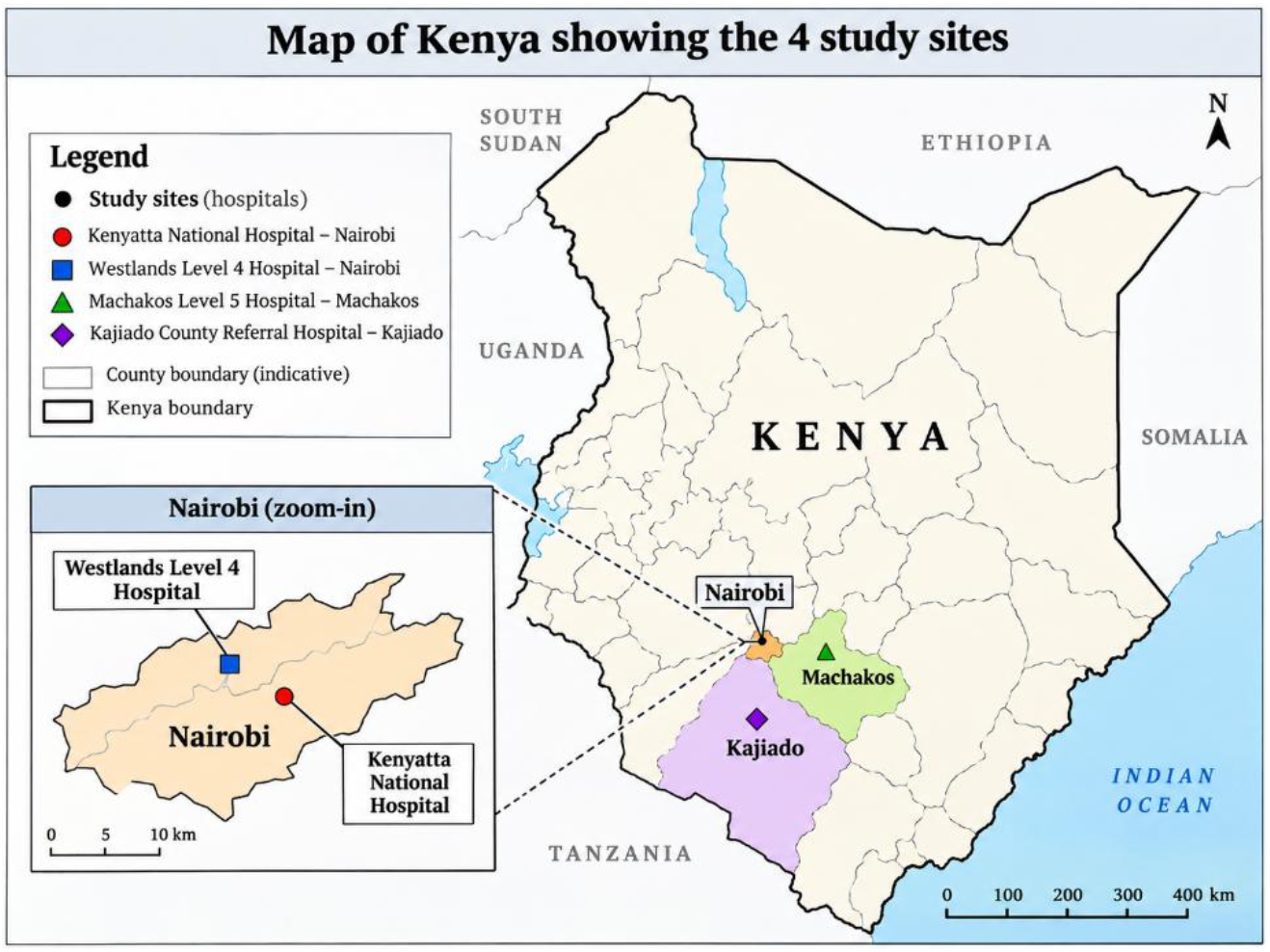
Map of Kenya showing the 4 study sites

### Study population, sample size and recruitment

Eligible participants were children, adolescents and young adults aged 1–34 years with clinician-documented type 1 diabetes. The participants had been in diabetes care for at least six months and were attending routine follow-up at one of the four participating clinics. Participants with acute or clinically significant illness, including severe hypoglycaemia, febrile or critical illness, or current hospital admission, were excluded to minimise transient alterations in thyroid-function tests related to acute metabolic or systemic illness. Pregnant participants and those using medications known to affect thyroid function or interfere with thyroid assays were also excluded because these factors may alter TSH and FT4 concentrations and complicate biochemical classification. Written informed consent was obtained from participants aged ≥18 years and from parents or guardians of minors, with age-appropriate assent obtained from children and adolescents where applicable.

The sample size was calculated using the single-population proportion formula, assuming a 14.7% prevalence of thyroid dysfunction, 95% confidence level and 5% precision. The minimum sample size was 193 and, after allowing for 10% non-response, increased to 212. A recruitment target of approximately 247 participants was set to improve precision and accommodate site variability. Participants were recruited consecutively during routine clinic visits at each study site. Recruitment targets were distributed across the four clinics according to their relative T1D patient volumes, such that higher-volume clinics contributed a larger proportion of participants. A total of 260 participants were enrolled: 92 from Kenyatta

National Hospital, 72 from Machakos Level 5 Hospital, 51 from Westlands Level 4 Hospital, and 45 from Kajiado County Referral Hospital. Of these, 244 had paired TSH and FT4 measurements and were included in the primary thyroid-function analysis, figure 2.

**Figure 2.**
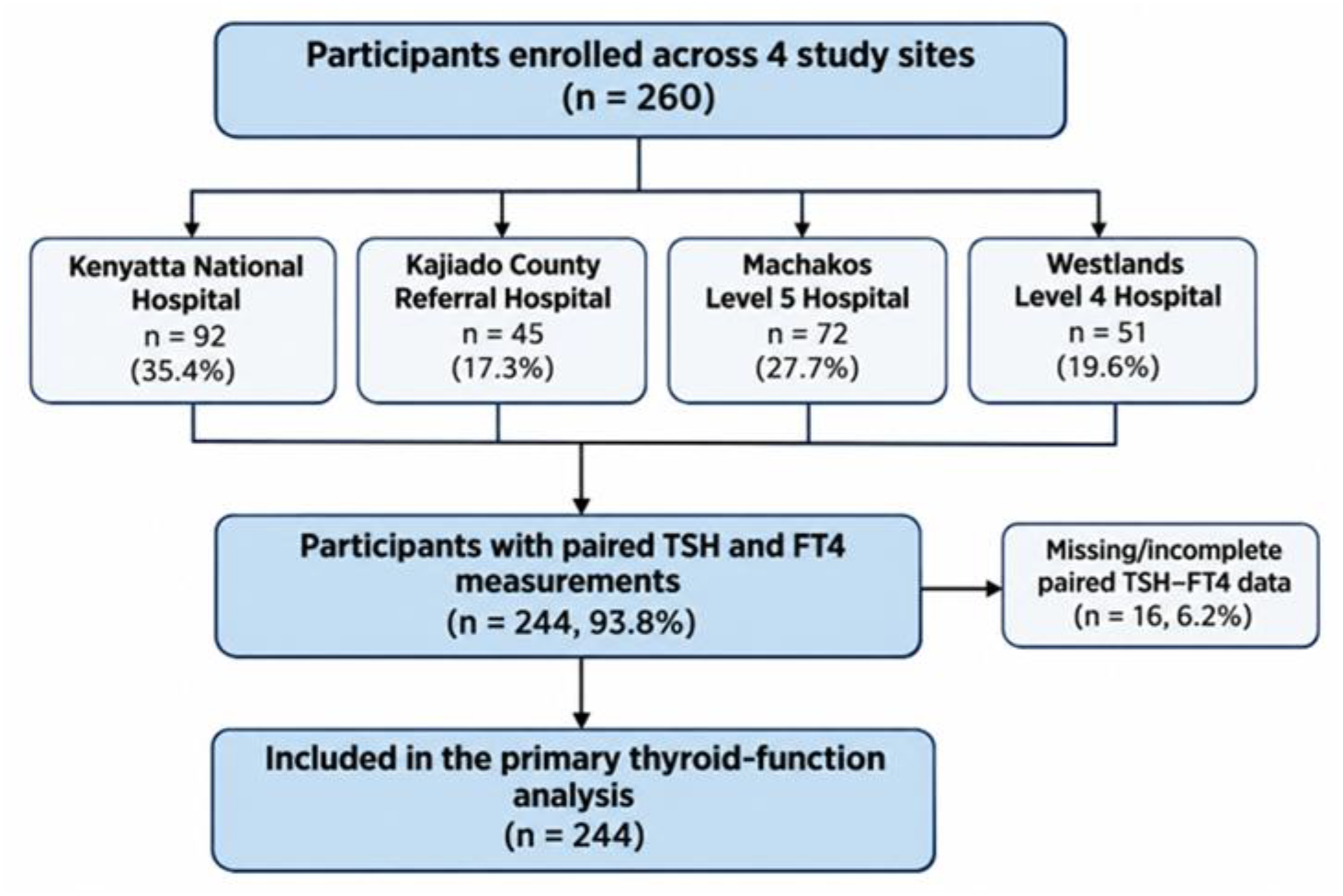
A flow diagram showing participant recruitment.

**Figure 3.**
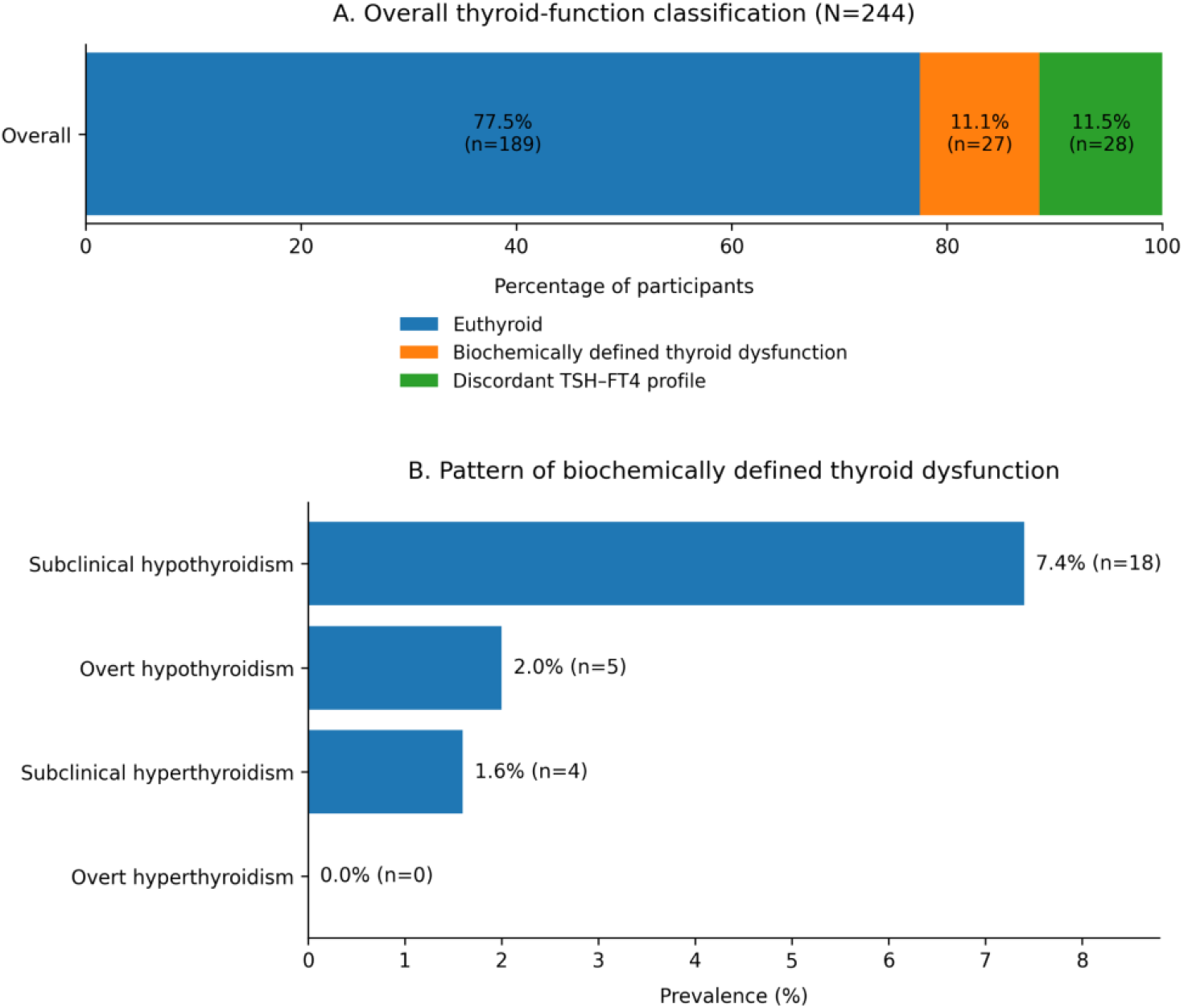
Prevalence and biochemical pattern of thyroid-function abnormalities among participants with type 1 diabetes (N=244). Panel A shows the overall distribution of euthyroidism, biochemically defined thyroid dysfunction and discordant TSH–FT4 profiles. Panel B shows the prevalence of individual thyroid dysfunction phenotypes.

Demographic and clinical information was obtained during the study visit and from the medical record, including age, sex, duration of T1D and glycated haemoglobin (HbA1c). Venous blood was collected during the study visit by trained phlebotomists. Around 5 cc of serum samples for thyroid-function testing were collected into serum-separator tubes, allowed to clot for approximately 30 minutes and centrifuged at approximately 1,500–2,000 × g for 10 minutes within two hours of collection. Serum was maintained at 2–8°C pending transport and transferred in temperature-controlled cool boxes to the central laboratory. All thyroid-function assays were performed centrally at Dr Kalebi Labs (DKL), an ISO 15189-accredited laboratory, to minimise inter-laboratory variation. Serum thyroid-stimulating hormone (TSH) and free thyroxine (FT4) were measured using Roche electrochemiluminescence immunoassays on the Roche cobas platform, according to manufacturer and laboratory standard operating procedures. The Roche cobas e 801 analytical unit uses electrochemiluminescence technology for immunochemistry assays and supports both TSH and FT4 testing. Internal quality control, assay-specific calibrators, equipment maintenance and external quality-assessment procedures were used before release of study results. The laboratory reference intervals used were TSH 0.51–4.17 mIU/L and FT4 12.6–21.0 pmol/L.

Thyroid-function status was determined from paired TSH and FT4 measurements. Participants were classified as euthyroid when both TSH and FT4 were within the laboratory reference intervals. Subclinical hypothyroidism was defined as TSH above the upper reference limit [>4.17 pmol/L] with FT4 within the reference range [12.6–21 pmol/L]; and overt hypothyroidism as elevated TSH [>4.17 pmol/L] with FT4 below the reference range [<12.6 pmol/L].^14,15^ Similarly, subclinical hyperthyroidism was defined as TSH below the lower reference limit [<0.51 pmol/L] with FT4 within the reference range [12.6–21 pmol/L]; and overt hyperthyroidism as low TSH [ <0.51 pmol/L] with FT4 above the reference range [>21 pmol/L].^16,17^ Participants with TSH–FT4 combinations that did not fulfil these conventional biochemical definitions were classified separately as having discordant thyroid-function profiles. These were henceforth excluded from the primary regression analyses comparing conventionally classifiable thyroid dysfunction with euthyroidism.

Participant characteristics were summarised using means and standard deviations or medians and interquartile ranges for continuous variables, as appropriate, and frequencies and percentages for categorical variables. The prevalence of euthyroidism, subclinical and overt hypothyroidism, subclinical and overt hyperthyroidism, and discordant thyroid-function profiles was calculated among participants with paired TSH and FT4 measurements, with 95% confidence intervals. Age and duration of T1D were additionally presented in categorical groups for descriptive analyses. Differences in categorical variables were assessed using Pearson’s χ^2^ or Fisher’s exact test, as appropriate. For association analyses, the primary outcome was conventionally classifiable thyroid dysfunction versus euthyroidism; participants with discordant thyroid-function profiles were excluded. Logistic regression was used to examine associations with age, sex and duration of T1D, with HbA1c included in an expanded model. Because hypothyroidism constituted the predominant thyroid phenotype, a prespecified secondary analysis compared participants with hypothyroidism with euthyroid participants. Odds ratios (ORs), adjusted odds ratios (aORs), 95% confidence intervals and two-sided p-values were reported, with statistical significance defined as p<0.05.

## Results

### 1. Participant characteristics

A total of 260 children, adolescents and young adults with T1D were included in the study, of whom 150 (57.7%) were female. Participants ranged in age from early childhood to 34 years, with the largest proportion aged 20–24 years. Nearly half of participants (48.8%) had lived with T1D for less than five years, while 29.2% had a diabetes duration of 5–9 years; only 2.3% had lived with T1D for ≥20 years. Glycaemic control was generally poor, with 248/257 (96.5%) participants having HbA1c ≥7%, table 1.

**Table 1:** Participant characteristics.

| **Characteristic** | **Overall** |
|---|---|
| **Age, years** | N=260 |
| Mean ± SD | **19.79 ± 8.02** |
| Median (IQR) | **21 (13)** |
| **Age group, n (%)** |  |
| 0–4 years | 7 (2.7) |
| 5–9 years | 24 (9.2) |
| 10–14 years | 45 (17.3) |
| 15–19 years | 39 (15.0) |
| 20–24 years | 66 (25.4) |
| 25–29 years | 47 (18.1) |
| 30–34 years | 32 (12.3) |
| **Sex, n (%)** |  |
| Female | **150 (57.7)** |
| Male | 110 (42.3) |
| **Duration of T1D, n (%)** |  |
| <5 years | **127 (48.8)** |
| 5–9 years | 76 (29.2) |
| 10–14 years | 36 (13.8) |
| 15–19 years | 15 (5.8) |
| ≥20 years | 6 (2.3) |
| **HbA1c, %,** N=257 |  |
| Mean ± SD | **11.36 ± 3.06** |
| Median (IQR) | 11.00 (3.55) |
| <7%, n (%) | 9 (3.5) |
| ≥7%, n (%) | **248 (96.5)** |
| **BMI, kg/m², N=260** |  |
| Mean ± SD | 21.22 ± 4.66 |
| Median (IQR) | 20.49 (6.00) |
| Low | 83 (31.9) |
| Normal | 138 (53.1) |
| Overweight | 39 (15.0) |
| **Insulin regimen, N=163** |  |
| Basal-bolus | 108 (66.3) |
| Premix | 55 (33.7) |

Among 244 participants with paired TSH and FT4 measurements, 189 (77.5%) were euthyroid, 27 (11.1%) had biochemically defined thyroid dysfunction, and 28 (11.5%) had discordant TSH–FT4 profiles. Subclinical hypothyroidism was the most common thyroid dysfunction phenotype (7.4%, n=18), followed by overt hypothyroidism (2.0%, n=5) and subclinical hyperthyroidism (1.6%, n=4); no overt hyperthyroidism was identified, figure 2.

The paired TSH–FT4 matrix demonstrated that most participants were euthyroid (n=189). The most frequent abnormal biochemical pattern was subclinical hypothyroidism (n=18), followed by overt hypothyroidism (n=5) and subclinical hyperthyroidism (n=4). Discordant profiles were mainly characterised by normal TSH with low FT4 (n=14) or elevated FT4 (n=12), with one participant each having low TSH with low FT4 and elevated TSH with elevated FT4, figure 4.

**Figure 4.**
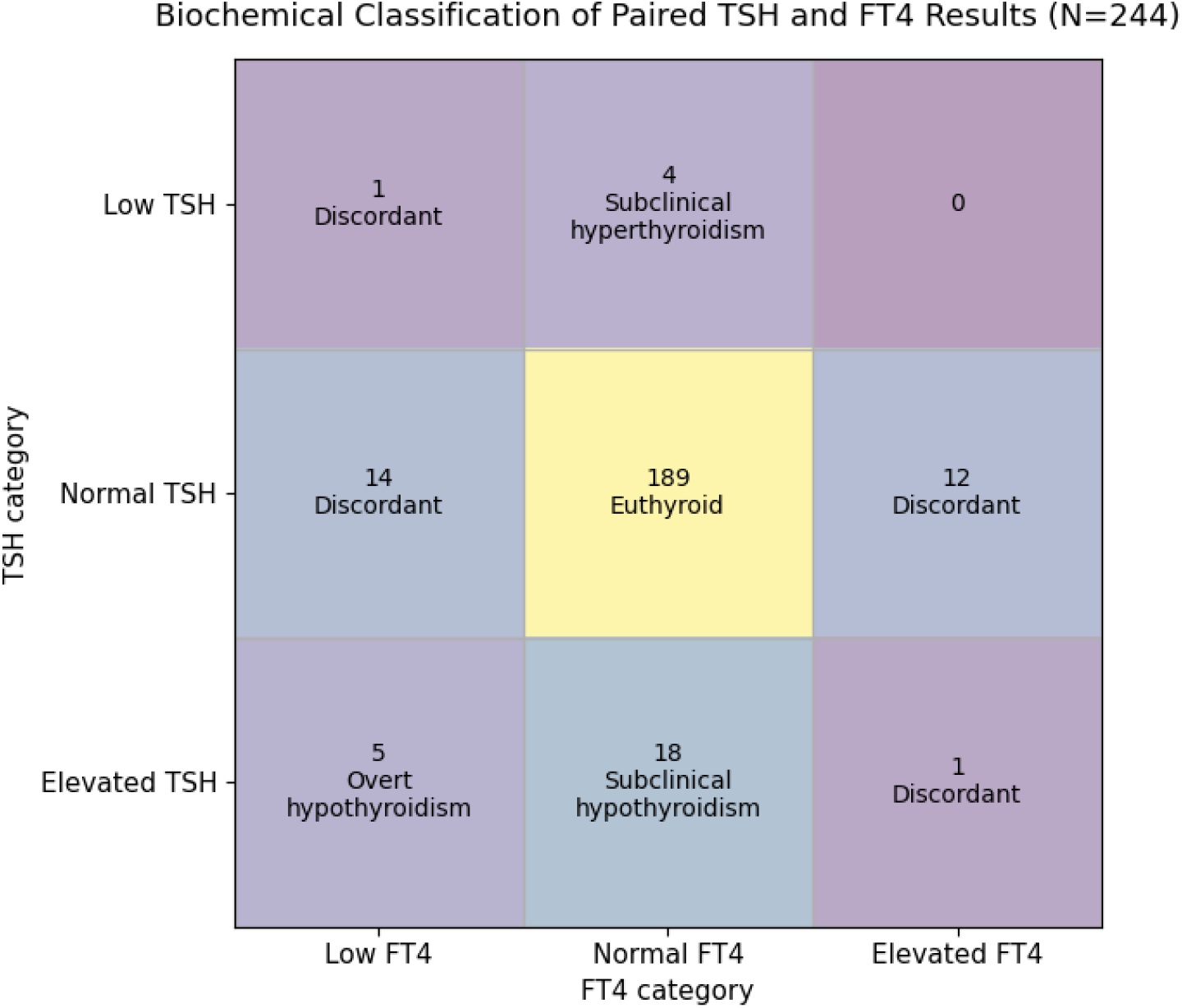
Biochemical classification of paired TSH and FT4 results among participants with type 1 diabetes (N=244). The matrix shows the distribution of participants across TSH and FT4 categories and the corresponding biochemical classification as euthyroid, subclinical or overt hypothyroidism, subclinical hyperthyroidism, or discordant TSH–FT4 profile.

Classifiable thyroid dysfunction was most frequent in the 10–14-year age group (17.5%), although there is no obvious progressive increase with age. It was more frequent among males than females (16.1% vs 9.8%). Similarly, no clear monotonic relationship with diabetes duration is apparent, table 2.

**Table 2.** Demographic and diabetes-related characteristics associated with classifiable thyroid dysfunction.

| Characteristic | Euthyroid, n (%) | Classifiable thyroid dysfunction, n (%) |
|---|---|---|
| **Age group** |  |  |
| 0–4 | 6 (85.7) | 1 (14.3) |
| 5–9 | 18 (85.7) | 3 (14.3) |
| 10–14 | 33 (82.5) | **7 (17.5)** |
| 15–19 | 27 (84.4) | 5 (15.6) |
| 20–24 | 47 (87.0) | 7 (13.0) |
| 25–29 | 35 (92.1) | 3 (7.9) |
| 30–34 | 23 (95.8) | 1 (4.2) |
| **Sex** |  |  |
| Female | 111 (90.2) | **12 (9.8)** |
| Male | 78 (83.9) | **15 (16.1)** |
| **Duration of T1D** |  |  |
| <5 years | 94 (88.7) | 12 (11.3) |
| 5–9 years | 54 (84.4) | **10 (15.6)** |
| 10–14 years | 23 (88.5) | 3 (11.5) |
| 15–19 years | 13 (92.9) | 1 (7.1) |
| ≥20 years | 5 (83.3) | 1 (16.7) |

Hypothyroidism was observed across all age groups but was proportionally most frequent among participants aged 10–14 years (17.5%). The prevalence was higher among males than females (15.2% vs 7.5%), although this difference did not reach statistical significance (p=0.073). No consistent trend was observed across increasing duration of diabetes, table 3.

**Table 3:** Hypothyroidism according to age, sex and duration of diabetes.

| Characteristic | Euthyroid | Hypothyroidism | Hypothyroidism within category |
|---|---|---|---|
| **Age group** |  |  |  |
| 0–4 | 6 | 1 | 14.3% |
| 5–9 | 18 | 3 | 14.3% |
| 10–14 | 33 | **7** | **17.5%** |
| 15–19 | 27 | 4 | 12.9% |
| 20–24 | 47 | 4 | 7.8% |
| 25–29 | 35 | 3 | 7.9% |
| 30–34 | 23 | 1 | 4.2% |
| **Sex** |  |  |  |
| Female | 111 | 9 | **7.5%** |
| Male | 78 | 14 | **15.2%** |
| **T1D duration** |  |  |  |
| <5 years | 94 | 12 | 11.3% |
| 5–9 years | 54 | 7 | 11.5% |
| 10–14 years | 23 | 2 | 8.0% |
| 15–19 years | 13 | 1 | 7.1% |
| ≥20 years | 5 | 1 | 16.7% |

In multivariable logistic regression, age and duration of T1D were not significantly associated with hypothyroidism. Male sex was associated with approximately twofold higher adjusted odds of hypothyroidism, although this did not reach statistical significance (aOR 2.13, 95% CI 0.87–5.20; p=0.098). Age (aOR 0.95, 95% CI 0.90–1.01; p=0.100) and duration of T1D (aOR 1.02, 95% CI 0.93–1.13; p=0.651) were also not statistically significant, figure 5.

**Figure 5.**
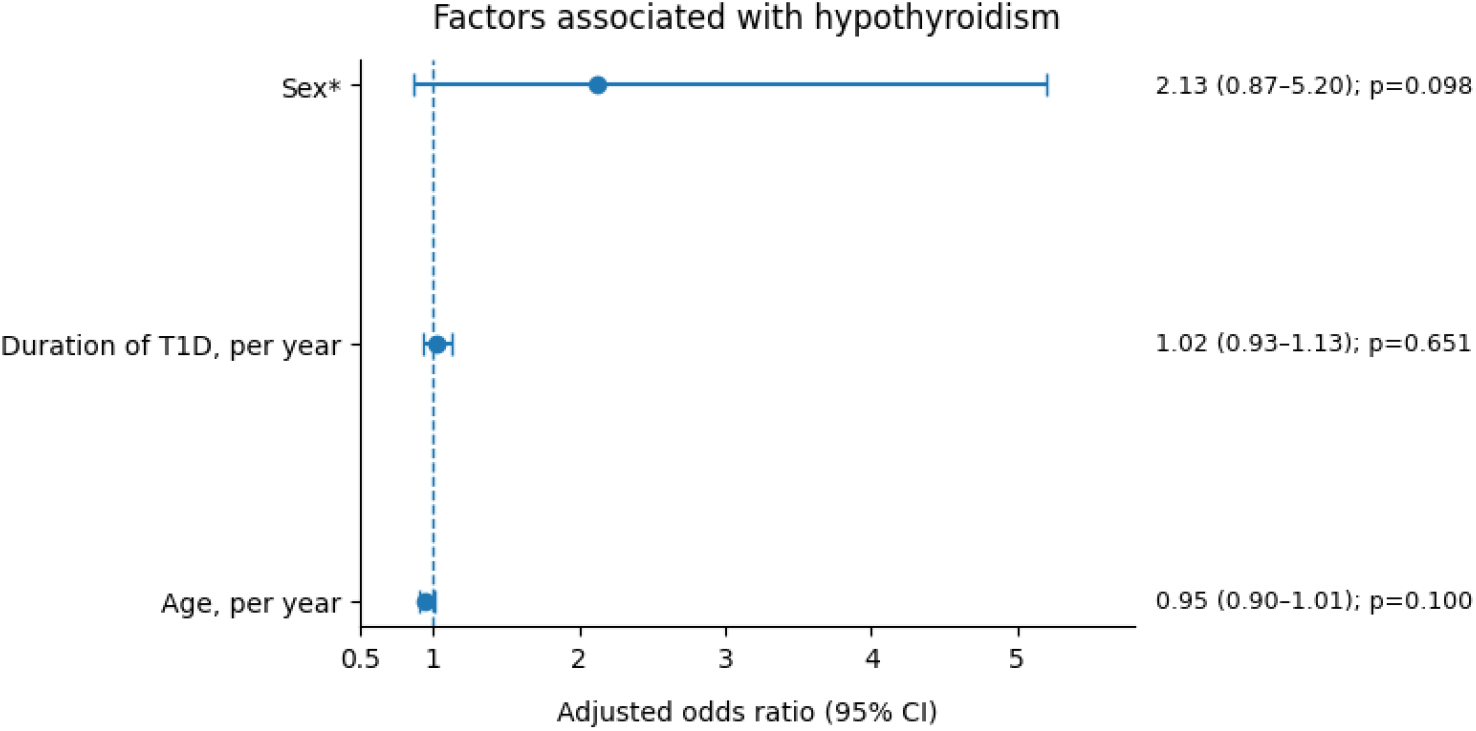
Factors associated with hypothyroidism among participants with type 1 diabetes. Points represent adjusted odds ratios and horizontal lines represent 95% confidence intervals from the multivariable logistic regression model. The vertical dashed line indicates the null value (aOR=1.0).

## DISCUSSION

In this multicentre Kenyan cohort of children, adolescents and young adults with type 1 diabetes, 11.1% had biochemically defined thyroid dysfunction. Hypothyroidism was the predominant abnormality, affecting 9.4% of participants, with subclinical hypothyroidism (7.4%) occurring more frequently than overt hypothyroidism (2.0%). Subclinical hyperthyroidism was uncommon (1.6%) and no overt hyperthyroidism was identified. An additional 11.5% had discordant TSH– FT4 profiles that did not fulfil the predefined biochemical criteria for hypothyroidism or hyperthyroidism. Age and duration of diabetes were not independently associated with hypothyroidism, while the observed higher odds among males did not reach statistical significance. These findings demonstrate a clinically relevant burden of predominantly subclinical thyroid dysfunction in young people with T1D in Kenya.

Hypothyroidism was the predominant thyroid abnormality in our cohort, affecting 9.4% of participants, with subclinical hypothyroidism (7.4%) more common than overt hypothyroidism (2.0%). This prevalence closely mirrors the pooled estimate of 9.8% reported in a meta-analysis of 293,889 individuals with T1D and the 9.9% reported among 19,225 children in the international SWEET registry.^4,5^ Regionally, our findings are somewhat higher than those from Uganda, where 7.3% of children and adolescents with T1D had subclinical hypothyroidism, and comparable with an Egyptian paediatric cohort reporting 6.3% subclinical and 2.1% overt hypothyroidism.^10,11^ The predominance of subclinical disease across these cohorts suggests that thyroid dysfunction in children and young people with T1D is frequently detected before overt biochemical disease develops. To our knowledge, contemporary multicentre Kenyan data specifically characterising TSH–FT4 abnormalities among children and young people with T1D remain very limited, underscoring the importance of the present study in providing locally relevant evidence on the burden and pattern of thyroid dysfunction.

A notable finding was that 11.5% of participants had discordant TSH–FT4 profiles, most commonly normal TSH with either low or elevated FT4. One possible explanation is non-thyroidal illness syndrome (NTIS), or euthyroid sick syndrome, in which metabolic stress alters peripheral thyroid-hormone metabolism and hypothalamic–pituitary–thyroid axis regulation, resulting in thyroid-function tests that do not conform to typical primary thyroid disease patterns.^18,19^ In children with T1D, poor metabolic control has been associated with lower T3, higher reverse T3 and other features of NTIS. Tahirović et al. demonstrated these abnormalities particularly among participants with HbA1c >10% and those with diabetic ketoacidosis.^20^ More recent paediatric DKA studies have similarly reported lower FT3, FT4 and TSH among children with euthyroid sick syndrome, with abnormalities related to higher glucose, HbA1c and severity of metabolic acidosis.^21-23^ Earlier studies of untreated T1D also documented normal TSH with low FT4, attributed partly to altered hypothalamic–pituitary regulation and peripheral thyroid-hormone metabolism.^23^ The ADA consequently cautions that thyroid-function tests may be misleading in the setting of marked hyperglycaemia, ketosis or ketoacidosis and recommends reassessment after metabolic stability.^1^ This may be relevant to our cohort, in which glycaemic control was generally poor, with a mean HbA1c of 11.36% and 96.5% of participants having HbA1c above the recommended target. Although participants with acute DKA were excluded, chronic metabolic dysregulation may have contributed to some of the discordant thyroid-function profiles observed. Other explanations, particularly for normal TSH with elevated FT4, include analytical or assay interference; persistent discordant results should therefore prompt repeat testing and, where necessary, assessment using an alternative assay platform.^19^

In multivariable analysis, age and duration of diabetes were not independently associated with hypothyroidism. Male participants had approximately twice the adjusted odds of hypothyroidism compared with females, although this did not reach statistical significance (aOR 2.13, 95% CI 0.87–5.20; p=0.098). This differs from the recent SWEET registry, where increasing age, female sex and longer diabetes duration were associated with hypothyroidism^5^ and from an Egyptian paediatric study reporting associations with older age and longer diabetes duration.^11^ However, other longitudinal data have shown thyroid-disease incidence to be relatively stable across age groups and independent of diabetes duration, although remaining higher in females.^3,4,24^ The absence of significant associations in our cohort may reflect the relatively small number of hypothyroidism cases, which limited statistical power to detect modest associations. In addition, the cross-sectional design captures thyroid status at a single point in time and therefore cannot assess how risk evolves with increasing age or duration of diabetes.

In this multicentre Kenyan cohort, thyroid dysfunction affected approximately one in nine children and young people with T1D and was predominantly hypothyroid, particularly subclinical hypothyroidism. Discordant thyroid-function profiles were also frequent, underscoring the need for careful biochemical interpretation and confirmation of abnormal results. Although both international guidance and the Kenya National Clinical Guidelines on Management of Diabetes Mellitus, 2024 recommend thyroid screening,^25,26^ consistent implementation across Kenyan diabetes clinics remains uncertain. Healthcare providers should strengthen routine thyroid surveillance within T1D care, while health-system leaders and policy makers should support its practical implementation through clear screening pathways and access to testing. Further longitudinal research is needed to determine the persistence, progression and clinical significance of both subclinical and discordant thyroid abnormalities in this population.

### Strengths and limitations

Strengths of this study include its multicentre design, broad age range and centralised TSH and FT4 testing using a single laboratory platform. Limitations include the cross-sectional design, single-point thyroid assessment and the small number of participants within individual thyroid-dysfunction categories, which reduced power for subgroup and regression analyses.

## Data Availability

All data produced in the present study are available upon reasonable request to the authors

## Statements and Declarations

### Ethical considerations

Ethical approval was obtained from the Kenyatta National Hospital–University of Nairobi Ethics and Research Committee (P291/04/2026). All study procedures were conducted in accordance with applicable ethical standards and regulations.

### Consent to participate

Written informed consent was obtained from all participants aged 18 years and older. For participants younger than 18 years, written informed consent was obtained from a parent or legal guardian, with age-appropriate assent obtained from the child or adolescent before enrolment.

### Consent for publication

Not applicable. The manuscript does not contain identifiable individual participant information, images, or videos.

### Contributors

PW conceived and drafted the manuscript. KM, MB, and PL, WS, AO critically reviewed the manuscript and contributed to its intellectual content. All authors reviewed and approved the final version.

## Acknowledgments

The authors gratefully acknowledge the children and young adults who participated in the study, as well as the clinical and laboratory teams at the participating study sites for their support during data and specimen collection.

## Use of generative artificial intelligence

A large language model was used during manuscript preparation to assist with improving sentence structure, and refining grammar and readability.

